# Nine-month all-oral regimen for MDR/RR-TB in Lesotho: safety and effectiveness by HIV

**DOI:** 10.64898/2026.09.04.26362253

**Authors:** Sara M. Sauer, Stephane Mpinda, Allison LaHood, Lawrence Oyewusi, Refiloe Maime, Joalane Makaka, Mahooe Thoakana, Daniel Monyaesa, Litsoanelo Musa, Tlotlisang Thai, Letizia Trevisi, Elna Osso, Phate Sebakeng, Meseret Tamirat, Marguerite Curtis, Andrew Lindeborg, Edmund Shen, Michael L. Rich, Carole D. Mitnick, Kwonjune Seung, Molly F. Franke, Mikanda K. Kunda, Llang Maama

## Abstract

**Background:** Individuals living with HIV have been under-represented in operational research and trials of all-oral regimens for multidrug- and rifampicin-resistant tuberculosis (MDR/RR-TB) treatment. We report outcomes from a prospective observational cohort study of a nine-month regimen comprised of bedaquiline, linezolid, a fluoroquinolone, delamanid, and clofazimine, in Lesotho, a setting with a high prevalence of HIV infection.

**Methods:** We analyzed patients who initiated the regimen between 2020 and 2023. Overall, and by HIV status, we characterized treatment safety by summarizing the frequency and timing of clinically relevant adverse events. We described treatment effectiveness at the end of treatment and sustained treatment success eighteen months after treatment initiation.

**Results:** Two hundred and fifty-five patients who initiated the shortened all-oral regimen were included in the analysis, of whom 161 (63.1%) were living with HIV. Regardless of HIV status, the most frequent clinically relevant adverse event was peripheral neuropathy, followed by myelosuppression. The estimated unadjusted rates of sustained treatment success at 18 months post-treatment initiation were 86.4% (95% confidence interval (CI): 81.4%, 90.5%) overall, 85.0% (95% CI: 78.2%, 90.4%) among patients living with HIV, and 88.8% (95% CI: 80.3%, 94.5%) among people without HIV.

**Conclusions:** The effectiveness of the nine-month all-oral regimen was excellent. Although adverse events were common, they were manageable and did not appear to substantially impact treatment success. Results were similar regardless of HIV status, supporting the use of this regimen in this population.

**Funding:** SMS was supported by the National Institutes of Allergy and Infectious Diseases under Award Number T32 AI007433. MK, AL, RM, JM, DM, LM, TT, LT, EO, MC, LO, MFF, CDM, AL, KS, and MLR were supported by the National Institutes of Allergy and Infectious Diseases under Award Number R01AI46095. The funders had no role in the conceptualization, analysis, or presentation of findings of this study.

## INTRODUCTION

Over the past decade, drugs such as bedaquiline, delamanid, linezolid, and clofazimine have been introduced or repurposed, enabling shorter and safer multidrug- and rifampicin-resistant tuberculosis (MDR/RR-TB) treatment regimens.^1,2^ In response to the growing body of evidence supporting the safety and effectiveness of shortened all-oral regimens,^3–5^ the World Health Organization (WHO) updated its MDR/RR-TB treatment guidelines in 2022 to prioritize the use of these regimens over longer and/or injectable-containing regimens.^1^ Specifically, the six-month BPaLM (bedaquiline, pretomanid, linezolid, moxifloxacin) regimen was to be used preferentially, with a nine-month seven-drug regimen proposed as a second option. In 2025, the WHO expanded these guidelines to include a second six-month regimen:^6^ the BEAT-Tuberculosis strategy involves the initiation of a five-drug regimen of bedaquiline, levofloxacin, linezolid, clofazimine, and delamanid, with subsequent tailoring dependent on the fluoroquinoline resistance testing results.^7^ If the TB isolate is found to be resistant, levofloxacin is discontinued; if susceptible, clofazimine is discontinued. If fluoroquinolone susceptibility testing is unavailable, as was the case in 18% of trial participants, all five drugs are retained. For instances in which an individual cannot receive a six-month regimen, the updated WHO guidance recommended three additional regimens, all tested in the endTB trial.^8^

While the 2025 WHO guidelines remarkably expand the options for all-oral shortened MDR/RR-TB treatments for fluoroquinolone-susceptible TB, the recommendations are conditional, due to the low certainty of evidence, and critical research gaps remain. For example, research on the safety and effectiveness of variations (i.e., with regard to duration, quantity, and composition) on currently recommended regimens is critical to ensuring treatment options are available to patients unable to take a recommended regimen (i.e., due to resistance or drug unavailability). A second outstanding question is whether regimen effectiveness varies across patient subgroups underrepresented in trials, potentially indicating closer monitoring and/or longer treatment for certain individuals. One such group is individuals living with HIV. Recent trials of shorter regimens have included too few people living with HIV to support confident subgroup inference. ^3,8^ It therefore remains unclear whether the safety and efficacy benefits observed overall extend to this population.

We analyzed data from STEM-TB (NCT05871489), a prospective observational cohort study that enrolled patients with fluoroquinolone-susceptible MDR/RR-TB in Lesotho, where the National Tuberculosis Program implemented a regimen comprising bedaquiline, levofloxacin, linezolid, delamanid, and clofazimine under operational research conditions. Although composed of the same medications as the BEAT-Tuberculosis initial regimen, it was intended to remain a five-drug regimen and was administered for nine months rather than six. We estimated safety and effectiveness overall and by HIV infection.

## METHODS

### Study setting

Lesotho has one of the world’s highest estimated TB incidence rates. WHO estimates that in 2020 (the year of study initiation) the TB incidence rate in Lesotho was 598.6 per 100,000 population, and that 3.9% and 7.5% of people with new and previously-treated pulmonary bacteriologically-confirmed TB, respectively, had MDR/RR-TB.^9^ Furthermore, an estimated 20.7% of adults aged 15-49 years were living with HIV in 2020.^10^

During the study period (2020-2023), all individuals initiating treatment for MDR/RR-TB in Lesotho did so at Botsabelo MDR-TB Hospital in Maseru, which was the national referral center for MDR-TB care in Lesotho. Treatment initiation occurred on an outpatient basis unless severe disease required hospitalization. After treatment initiation, clinical management remained under the supervision of Botsabelo staff, who conducted regular monthly follow-up visits in patients’ home districts throughout the course of therapy. Although MDR/RR-TB services began to be decentralized to one district-level facility during the study period, only a small number of patients were formally transferred from Botsabelo to receive ongoing care at that site.

### Study population

From September 2020 to October 2023, we prospectively enrolled individuals with bacteriologically-confirmed MDR/RR-TB initiating the standardized nine-month all oral regimen. Patients were not eligible for treatment with the shorter regimen if they met any of the following exclusion criteria: not eligible to take a fluoroquinolone due to resistance, prior exposure of at least one month, or allergy/intolerance; evidence of resistance, prior exposure of at least one month or known allergy to two or more of the drugs included in the regimen; extrapulmonary disease without pulmonary involvement; baseline AST or ALT more than 5 times the upper limit of normal; taking any medications contraindicated with the drugs in the shorter MDR-TB regimen; unable to attend or comply with treatment or follow-up schedule; unable to take oral medication; taking part in a clinical trial of any medicinal product; had a baseline QTcF interval of more than 500 ms; or any condition that, based on clinical discretion, would preclude treatment with a standardized treatment, such as severe forms of extrapulmonary TB.

### Standardized all-oral treatment regimen

The National Tuberculosis and Leprosy Program determined the composition of the all-oral shortened MDR/RR-TB treatment based on patient characteristics, prevailing drug resistance profiles, and the intensity of anti-TB drug use over the preceding decade. The study regimen comprised 39 weeks of bedaquiline (400 mg once daily for the first 2 weeks, followed by 200 mg three times weekly for the remainder of the 39 weeks), linezolid (600 mg daily), levofloxacin, clofazimine, and delamanid. Drugs could be reintroduced after an interruption, at the discretion of the clinician. For linezolid, reintroduction could occur at the standard dose of 600 mg daily or at a reduced dose (e.g. 300 mg daily or 600 mg three times weekly). Depending on treatment history and baseline drug susceptibility testing (DST), participants were eligible to initiate treatment with only four of the five study agents, provided a fluoroquinolone was used.

Medications were directly observed throughout treatment, relying on treatment supporters. Treatment could be extended up to 12 months based on clinical discretion and considering microbiologic and radiographic evolution after four months of therapy, the presence of pulmonary lesions equal or larger than two lung fields, or other comorbidities, including HIV and diabetes mellitus.

### Study procedures

The endTB clinical treatment guide for all-oral shortened treatment regimens informed patient monitoring.^11^ Baseline evaluation included a thorough medical history, clinical exam, pregnancy testing, sputum collection, and blood collection for laboratory tests, chest X-ray, and ECG . Peripheral blood testing included a complete blood count, hemoglobin, creatinine/urea, electrolytes, albumin, liver function tests, and HIV, hepatitis B, and hepatitis C screening.

Laboratory evaluations performed on the collected sputum sample included smear microscopy, culture, Xpert MTB/RIF (or other molecular resistance testing, as available), and phenotypic first and second-line DST. Resistance testing to bedaquiline and delamanid was not conducted.

Follow-up smear and culture were conducted monthly throughout treatment. Routine follow-up visits included a review of signs and symptoms and a clinical exam, which covered visual acuity and color differentiation (assessed using a Snellen Chart and Ishihara Color Plates, respectively), and screened for peripheral neuropathy using the symptom-based ACTG Brief Peripheral Neuropathy Screen.^12^ A peripheral neuropathy diagnosis required a subjective neuropathy symptom score >0 and at least one abnormal bilateral objective finding (vibratory sense or deep tendon ankle reflex). The subjective sensory score was used to assign adverse event grade. Monthly routine testing consisted of an ECG, complete blood count, creatinine/urea, serum electrolytes, and liver function tests. Adverse events were reported and graded using the MSF Severity Grading Scale;^13^ pre-defined thresholds (Table 1) were used to determine clinical relevance. Following treatment completion, patients were advised to return monthly for 12 months of post-treatment follow-up. Sputum for smear and culture was collected at each visit. A monthly food ration for a family of four was provided throughout treatment and the twelve months of post-treatment follow-up.

**Table 1.** Adverse Events of Special Interest and Severity Scale Threshold for Clinically Relevant Adverse Events of Special Interest: STEM-TB Cohort.

| **AESI** | **Severity Scale or “Other” Terms Included** | **Grade(s)** | **Definition for Minimum Grade** |
| --- | --- | --- | --- |
| Hepatotoxicity | Alanine aminotransferase (ALT) increased | ≥3 | Grade 3: alanine aminotransferase and/or aspartate aminotransferase >5 times the upper limit of normal |
|  | Aspartate aminotransferase (AST) increased | ... | ... |
|  | Hepatotoxicity | ... | ... |
|  | Hepatitis | ... | ... |
| Myelosuppression | Anemia | Anemia ≥3 | Grade 3: hemoglobin <7.9 g/dL |
|  | Platelets decreased | Thrombocytopenia ≥3 | Grade 3: platelets decreased <50 000/mm<sup>3</sup> |
|  | White blood cells decreased | Leukopenia ≥3 | Grade 3: white blood cells decreased <2000/mm<sup>3</sup> |
|  | Lymphocyte count decreased | Lymphocytopenia ≥3 | Grade 3: lymphocytes decreased <500/mm<sup>3</sup> |
|  | Absolute neutrophil count | Neutropenia ≥2 | Grade 2: absolute neutrophil count <750/mm<sup>3</sup> |
|  | Pancytopenia | Pancytopenia ≥2 | Grade 2: any combination of the above |
| Optic neuritis | Optic nerve disorder (optic neuritis) | All grades | Grade 1: clinical diagnosis, no symptoms |
| Peripheral neuropathy | Neurosensory disorders | All grades | Grade 1: mild impairment or discomfort/Brief Peripheral Neuropathy Screen (BPNS) subjective sensory neuropathy score 1–3 on any side |
|  | Paresthesia (burning, tingling, etc) | ... | ... |
| QTc prolongation | Electrocardiogram<br>QT-corrected interval<br>prolonged | ≥3 | Grade 3: QTc ≥501<br>ms, no symptoms |

Table 1 has been reproduced with permission from Rashitov et al., *Clinical Infectious Diseases* 2024;79(4):1046–53, under the terms of the Creative Commons Attribution-Non Commercial-No Derivatives (CC BY-NC-ND) license.

### Sample size

Because the study was descriptive in nature, no formal sample size calculation was conducted. Instead, all consecutive individuals initiating treatment at during the study period were included.

### Statistical analysis

<u>Safety.</u> To characterize the safety of the shortened all-oral nine-month regimen, we considered five clinically relevant adverse events, namely hepatotoxicity, myelosuppression, optic neuropathy, peripheral neuropathy, and QTc prolongation, all known to be associated with the drugs used in the shortened regimen. The severity scale thresholds for the five clinically relevant adverse events of special interest are shown in Table 1. For each clinically relevant adverse event occurring on treatment, we computed its frequency, the median time to the first occurrence, and the incidence of the first event per 1000 person-months, overall and by HIV infection status.

Frequency was computed as the percentage of patients who experienced the specified adverse event at least once during treatment. Person-months of follow-up were calculated separately for each adverse event from the start of treatment to the date of the first occurrence of the adverse event (or to the end of treatment if the adverse event did not occur). Individuals who were enrolled but subsequently excluded from the study (i.e., due to late detection of baseline drug resistance) contributed person-time from treatment initiation until exclusion or the date of the adverse event, whichever came first. For each drug in the regimen, we also computed the percentage of patients with permanent drug discontinuation prior to 244 days.

<u>Effectiveness.</u> To assess regimen effectiveness, we describe outcomes at the end of treatment, after 18 months (corresponding to the follow-up of most trials of shortened treatment), and at the end of follow-up (21 months post-treatment initiation) overall and by HIV status, among patients who were not excluded. End-of-treatment outcomes were classified by clinicians into one of the following categories based on WHO definitions: cure, completion, death, failure of treatment, loss to follow-up (LTFU), or not evaluated.^14,15^ Under these definitions, an outcome of cure can only be assigned to an individual with bacteriologically-confirmed TB; all individuals in the cohort met this criterion. We estimated end-of-treatment success as the frequency of patients cured or completed treatment, excluding the small number who were not evaluated due to transfer out of the program (to South Africa).While excluding those not evaluated represents a departure from the WHO definition of treatment success, this definition better aligned with the research objective to evaluate a specific regimen rather than an overall program; not only was it unlikely that those treated outside of Lesotho would have received the regimen of interest, the composition and duration of their regimen could not be verified. Among individuals with treatment success, we estimated the frequency of recurrent TB and post-treatment death in the 12 months post-treatment completion (i.e. the end of follow-up), excluding patients LTFU post-treatment. We also calculated rates of recurrent TB and post-treatment death in the twelve months post-treatment completion by dividing the number of events by the sum of the follow-up months among those with treatment success. For the rate calculation, patients who were LTFU in the post-treatment period contributed to person time while they were under follow-up.

To estimate sustained treatment success 18 months after treatment initiation, we calculated the prevalence of a binary endpoint indicating a favorable versus unfavorable outcome at that time. Patients were classified as having an unfavorable outcome if they experienced any of the following before 18 months post-treatment start: 1) any-cause death during or after treatment, 2) loss to follow-up during treatment, 3) treatment failure, 4) recurrence after successful treatment. All other patients were classified as having experienced a favorable outcome by 18 months.

Consistent with the primary analysis, the small number of patients who were not evaluated in either the treatment or post-treatment period (i.e., transferred out during treatment or were lost to follow-up after treatment ended) were excluded). We examined whether end-of-treatment or final treatment outcomes varied by confirmed fluoroquinolone susceptibility, versus absence of a result. We also conducted a sensitivity analysis in which we recalculated effectiveness, considering only post-treatment deaths related to TB.

This study was approved by the Lesotho Ministry of Health National Ethics Committee and the Harvard Longwood Campus Institutional Review Board. All participants gave written informed consent.

## RESULTS

Of 331 individuals screened for inclusion, 76 (23%) were excluded, most commonly due to prior treatment with second-line drugs or fluoroquinolone resistance (n=34) or clinical discretion (n=26); remaining reasons are detailed in Figure 1.Two hundred and fifty-five participants were enrolled, of whom 161 (63.1%) were living with HIV. Table 2 shows the baseline characteristics: 30.2% of participants were female, the median age was 41 years (IQR: 31, 55), 57.3% were employed, and 38.2% were self-described smokers. Furthermore, 8.3% of patients had diagnosed diabetes, 5.3% had hepatitis B, 2.8% had hepatitis C, 37.8% had low BMI, 56.9% had anemia, 45.7% had bilateral disease, and 31.2% had cavitary disease. The majority (55.5%) of patients had no history of TB treatment, while 44.1% had been treated with first-line drugs only, and 0.4% with second-line drugs. Among people living with HIV, the median CD4 count was 242.5 cells/mm^3^ (IQR: 83.5 cells/mm^3^, 527.0 cells/mm^3^), and 88.1% were on antiretroviral treatment with >95% receiving a dolutegravir-based regimen.

**Figure 1.**
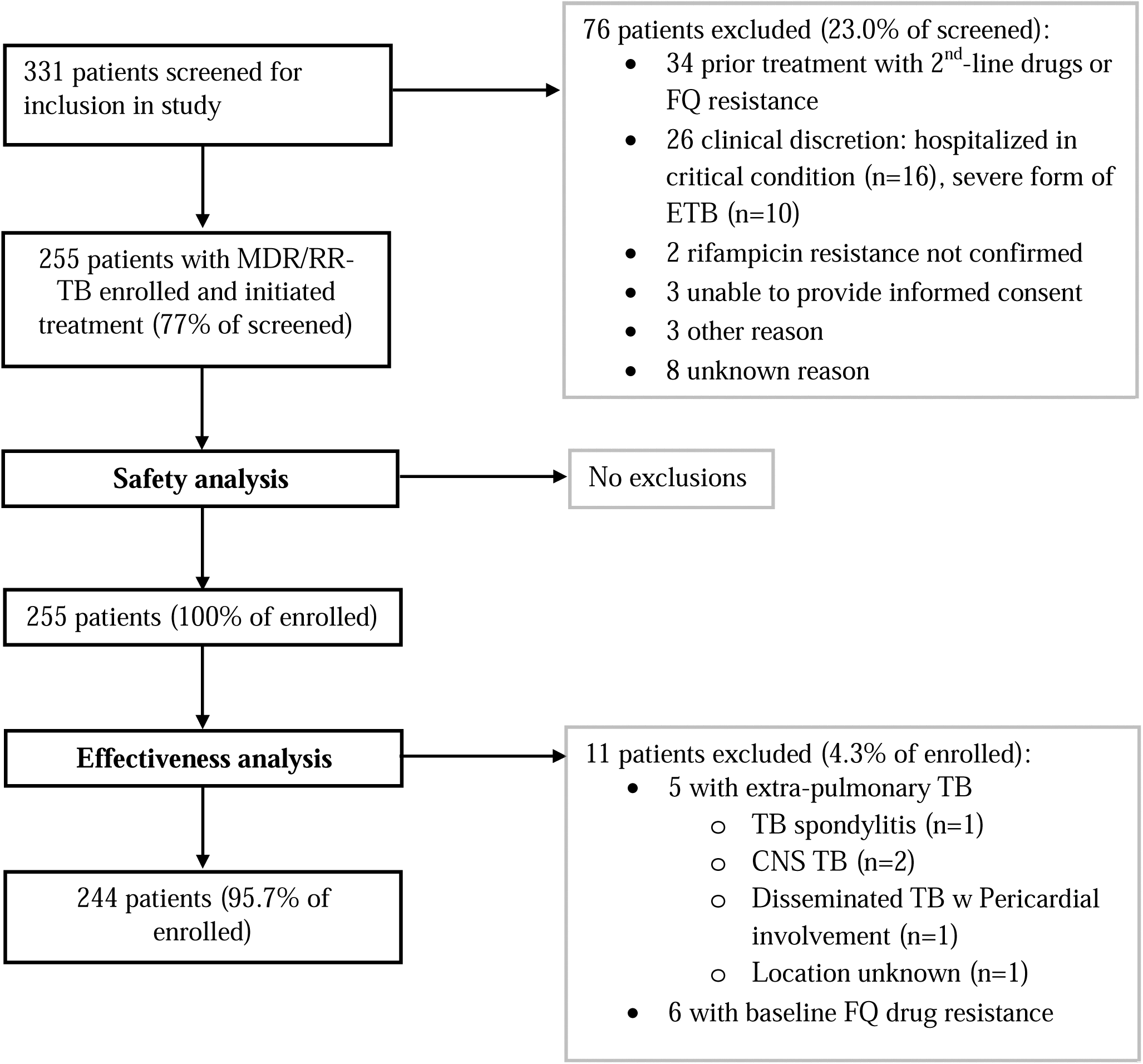
Flowchart of patients initiating a shortened all-oral regimen for multidrug-resistant or rifampin-resistant tuberculosis in Lesotho, STEM-TB cohort

**Table 2:** Baseline characteristics of patients who received shortened all-oral regimen for MDR/RR-TB in Lesotho, overall and by HIV status.

| Characteristic | N | Overall<br>N = 255<sup>†</sup> | HIV<br>N = 161<sup>†</sup> | No HIV<br>N = 94<sup>†</sup> |
| --- | --- | --- | --- | --- |
| Age | 255 | 41.0 (31.0, 55.0) | 40.0 (34.0, 49.0) | 47.5 (27.0, 64.0) |
| Female | 255 | 77 (30.2%) | 51 (31.7%) | 26 (27.7%) |
| Employed | 255 | 146 (57.3%) | 90 (55.9%) | 56 (59.6%) |
| Married or living together | 228 | 118 (51.8%) | 71 (50.4%) | 47 (54.0%) |
| Uses alcohol | 253 | 133 (52.6%) | 88 (55.3%) | 45 (47.9%) |
| Smoker | 254 | 97 (38.2%) | 64 (40.0%) | 33 (35.1%) |
| History of incarceration | 255 | 35 (13.7%) | 26 (16.1%) | 9 (9.6%) |
| Anemia | 255 | 145 (56.9%) | 102 (63.4%) | 43 (45.7%) |
| Malnutrition (BMI < 18.5) | 254 | 96 (37.8%) | 60 (37.5%) | 36 (38.3%) |
| Diabetes mellitus | 252 | 21 (8.3%) | 9 (5.7%) | 12 (12.9%) |
| Hepatitis B | 245 | 13 (5.3%) | 12 (7.9%) | 1 (1.1%) |
| Hepatitis C | 247 | 7 (2.8%) | 3 (1.9%) | 4 (4.4%) |
| TB treatment history | 254 |  |  |  |
| No prior TB treatment |  | 141 (55.5%) | 84 (52.5%) | 57 (60.6%) |
| First line drugs only |  | 112 (44.1%) | 75 (46.9%) | 37 (39.4%) |
| Second line drugs |  | 1 (0.4%) | 1 (0.6%) | 0 (0.0%) |
| Bilateral disease | 221 | 101 (45.7%) | 59 (43.1%) | 42 (50.0%) |
| Cavitary disease | 221 | 69 (31.2%) | 44 (32.1%) | 25 (29.8%) |
| Culture positive | 246 | 170 (69.1%) | 112 (72.7%) | 58 (63.0%) |
| Smear | 243 |  |  |  |
| □□□□ Negative |  | 127 (52.3%) | 78 (51.0%) | 49 (54.4%) |
| □□□□ Scanty |  | 11 (4.5%) | 4 (2.6%) | 7 (7.8%) |
| □□□□ One plus |  | 25 (10.3%) | 20 (13.1%) | 5 (5.6%) |
| □□□□ Two plus |  | 18 (7.4%) | 11 (7.2%) | 7 (7.8%) |
| □□□□ Three plus |  | 62 (25.5%) | 40 (26.1%) | 22 (24.4%) |
| Cavitary disease and smear of two plus or three plus | 211 | 37 (17.5%) | 24 (18.3%) | 13 (16.3%) |
| CD4 count | 160 | -- | 242.5 (83.5, | -- |
|  |  |  | 527.0) |  |
| On ART | 160 | -- | 141 (88.1%) | -- |
| Baseline ART regimen<br>Dolutegravir-based*<br>Other | 141 |  | 135 (95.7%)<br>9 (4.3) |  |
| <sup>†</sup>Median (Q1, Q3); n (%)<br>*132 participants received dolutegravir, lamivudine and tenofovir disoproxil fumarate; 3 received dolutegravir, lamivudine and abacavir |  |  |  |  |

Of the 255 patients who were initiated on the nine-month all-oral regimen, 11 (4.3%) were late exclusions due to baseline drug resistance (n=6) or based on clinical discretion due to severe extra-pulmonary disease in addition to pulmonary TB (n=5) (see Figure 1). All of these late exclusions occurred in the first few weeks of treatment as soon as the extrapulmonary TB was diagnosed or the DST result became available.

<u>Safety.</u> One hundred eighty-seven (73.3%) patients experienced at least one of the five clinically relevant adverse events of special interest. The most common clinically relevant adverse event was peripheral neuropathy, followed by myelosuppression, with 52.6% and 29.4% of patients experiencing these events at least once, respectively (Table 3). The frequency and incidence rate of myelosuppression and optic neuritis tended to be higher among patients living with HIV, while the frequencies and incidence rates of hepatotoxicity, peripheral neuropathy, and QTc prolongation tended to be higher among patients without HIV.

**Table 3:** Timing and frequency of clinically-relevant adverse events of special interest among patients receiving a shortened regimen for MDR/RR-TB in Lesotho, overall and by HIV status.

| **Clinically Relevant Adverse Event** | **Overall**<br>**(N=255)** | **HIV**<br>**(N=161)** | **No HIV**<br>**(N=94)** | **P-value\*** |
| --- | --- | --- | --- | --- |
| Hepatotoxicity<br>*Frequency, n (%)*<br>*Months to first clinically relevant AESI, median (IQR)*<br>*Rate /1000 person-months (95% CI)* | 50 (19.6)<br>2.4 (0.5, 5.4)<br>23.8<br>(17.7, 31.4) | 31 (19.2)<br>1.1 (0.5, 4.6)<br>23.4<br>(15.9, 33.1) | 19 (20.2)<br>3.4 (1.2, 6.1)<br>24.6<br>(14.8, 38.4) | 0.884 |
| Myelosuppression<br>*Frequency, n (%)*<br>*Months to first clinically relevant AESI, median (IQR)*<br>*Rate /1000 person-months (95% CI)* | 75 (29.4)<br>3.6 (1.9, 4.5)<br>38.9<br>(30.6, 48.7) | 53 (32.9)<br>2.9 (1.3, 4.6)<br>45.0<br>(33.7, 58.9) | 22 (23.4)<br>3.9 (2.5, 4.5)<br>29.2<br>(18.3, 44.3) | 0.097 |
| Optic neuritis<br>*Frequency, n (%)*<br>*Months to first clinically relevant AESI, median (IQR)*<br>*Rate /1000 person-months (95% CI)* | 3 (1.2)<br>2.6 (1.7, 2.7)<br>1.3<br>(0.3, 3.8) | 3 (1.9)<br>2.6 (1.7, 2.7)<br>2.0<br>(0.4, 5.9) | 0 (0.0)<br>--<br>0<br>(0.0, 4.3) | 0.302 |
| Peripheral neuropathy<br>*Frequency, n (%)*<br>*Months to first clinically relevant AESI, median (IQR)*<br>*Rate /1000 person-months (95% CI)* | 134 (52.6)<br>4.6 (3.4, 6.3)<br>76.2<br>(63.8, 90.2) | 77 (47.8)<br>4.6 (3.5, 6.0)<br>68.1<br>(53.7, 85.1) | 57 (60.6)<br>4.7 (3.3, 6.4)<br>90.8<br>(68.8, 117.7) | 0.105 |
| QTc prolongation<br>*Frequency, n (%)*<br>*Months to first clinically relevant AESI, median (IQR)* | 13 (5.1)<br>6.3 (3.0, 8.3) | 5 (3.1)<br>7.3 (3.6, 8.3) | 8 (8.5)<br>5.4 (2.4, 6.9) |  |
| *Rate /1000 person-months*<br>*(95% CI)* | 5.6<br>(3.0, 9.6) | 3.4<br>(1.1, 7.8) | 9.7<br>(4.2, 19.0) | 0.078 |

\* p-value comparing the rate of each adverse event by HIV status calculated using the exact Poisson method

<u>Regimen modifications.</u> Clinician-directed modifications to the standardized regimen occurred, most frequently with regard to the use of linezolid and selection of the fluoroquinolone. Discounting the 11 late exclusions, 15 of 244 individuals (6.2%) never initiated linezolid (7.2% of those living with HIV and 4.3% of those without HIV), and in 10 additional individuals (4.1%), linezolid was initiated >15 days after treatment initiation (median days: 55.5; interquartile range: 27–80). Eleven participants (4.5%) initiated a regimen with moxifloxacin instead of levofloxacin, and among the 233 who received levofloxacin, 15 (6.4%) were subsequently switched to moxifloxacin for >30 days. Additionally, 2 individuals received >30 days of cycloserine (31 and 211 days), 1 received 87 days of pyrazinamide, and 1 received 37 days of isoniazid.

Omitting 11 participants who were excluded late, 72 of 229 individuals who initiated linezolid (31.4%) had the dose reduced, and permanent drug discontinuation occurred in 3 of 244 individuals (1.2%) for bedaquiline, 15 of 233 (6.4%) for levofloxacin, 138 of 229 (60.3%) for linezolid, 3 of 243 (1.2%) for clofazimine, and 7 of 244 (2.9%) for delamanid (Table 4), with 61.2% of individuals having at least one drug permanently discontinued. The median duration of linezolid among those in whom it was permanently discontinued was 3.6 months (IQR: 1.7 to 4.8). Individuals living with HIV did not experience an elevated rate of linezolid discontinuation relative to those without HIV (83/141 (58.9%) and 55/88 (62.5%), respectively).

**Table 4.** Permanent drug discontinuation among patients who initiated a shortened all-oral regimen for multidrug-resistant or rifampin-resistant tuberculosis in Lesotho, STEM-TB cohort (N=244)

| **Drug (N who initiated the drug)** | **Permanently discontinued**<br>**All participants**<br>**n (%)** |
| --- | --- |
| Bedaquiline (N=244) | 3 (1.2) |
| Linezolid (N=229) | 138 (60.3) |
| Levofloxacin (N=233) | 15 (6.4) |
| Delamanid (N=244) | 7 (2.9) |
| Clofazimine (N=243) | 3 (1.2) |

<u>Effectiveness - end of treatment outcomes.</u> The median treatment duration was 39.9 weeks (IQR 39.0, 41.0). Two hundred and sixteen of 244 patients were cured (88.5%), while 19 (7.8%) died, 3 (1.2%) experienced treatment failure, 2 (0.8%) were LTFU during treatment, and 4 (1.6%) were not evaluated because they transferred their care to South Africa (Table 5). One treatment failed due to prolonged lack of culture conversion, another to bacteriologic reversion after initial culture conversion, and the third due to prolonged clinical deterioration despite initial culture conversion. Among the 19 deaths, the median time-to-death was 56.0 days (IQR 47.5-110.5), and all deaths were classified as TB-related. Two deaths were considered related and 3 were considered possibly-related to TB drugs.

**Table 5:**
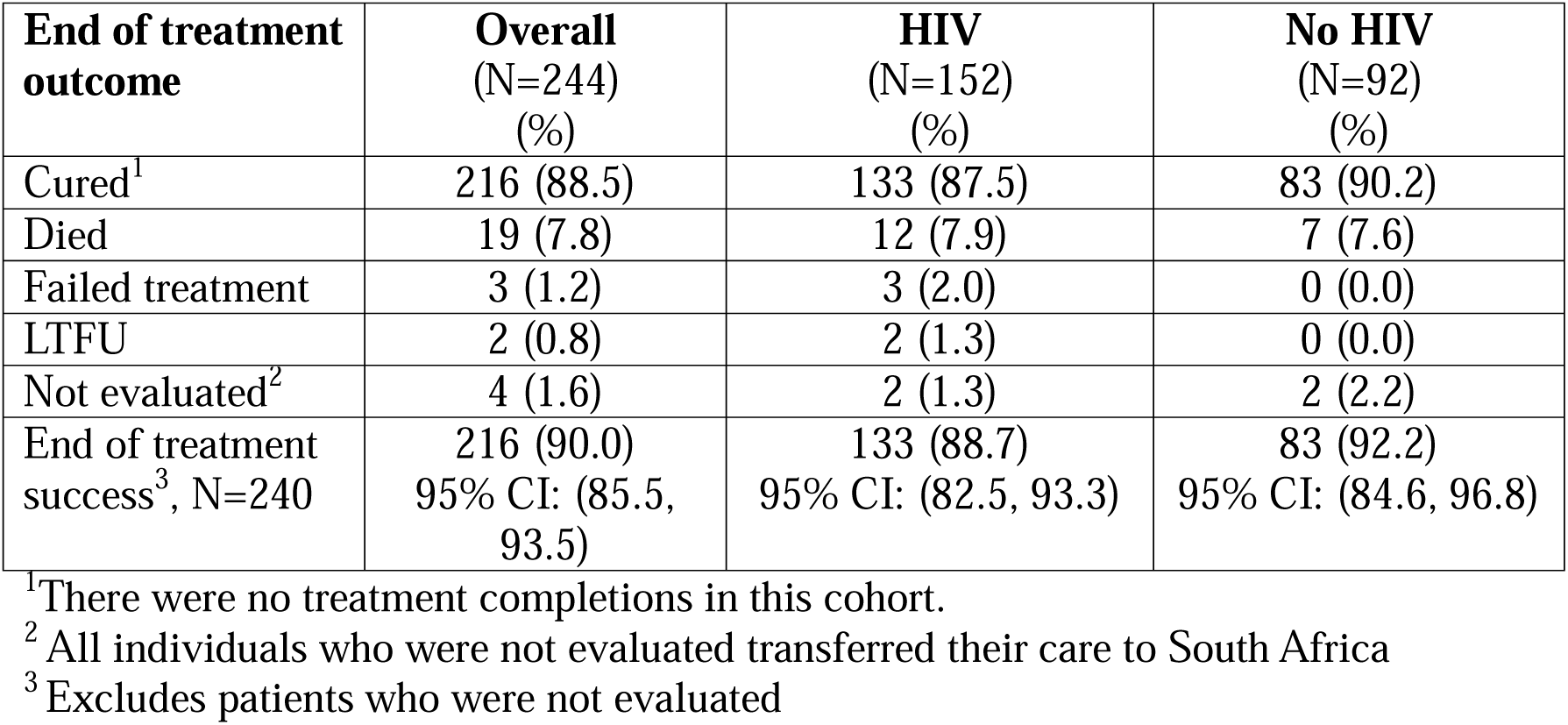
End-of-treatment outcomes among patients receiving a shortened regimen for MDR/RR-TB in Lesotho, overall and by HIV status.

The overall end-of-treatment success rate, excluding those not evaluated, was 90.0% (95% confidence interval (CI): 85.5%, 93.5%). Among patients living with HIV, the end of treatment success rate was 88.7% (95% CI: 82.5%, 93.3%), and among patients without HIV it was 92.2% (95% CI: 84.6%, 96.8%). The percentage of patients without a fluoroquinolone resistance test result was 47.5%. End of treatment results were similar among those without a fluoroquinolone test result, with an end-of-treatment success rate of 88.6% (95% CI: 81.3%, 93.8%) overall, 87.0% (95% CI: 76.7%, 93.9%) among individuals with HIV, and 91.1% (95% CI: 78.8%, 97.5%) among individuals without HIV (Table S2 in the Supporting Information).

<u>Effectiveness - post treatment outcomes.</u> Table 6 shows the distribution of the 18-month effectiveness outcomes. Overall, 204 (83.2%) of patients experienced a favorable outcome within 18 months post-treatment initiation, while 33 (13.1%) experienced an unfavorable outcome, and 8 (3.3%) were not evaluated. Among patients who experienced cure, 1 death related to TB, 3 deaths unrelated to TB, and 4 cases of recurrent TB were observed. Among patients with HIV, 125 (82.2%) had a favorable outcome at 18 months, 22 (14.5%) had an unfavorable outcome, and 5 (3.3%) were not evaluated, and among patients without HIV, 79 (85.9%) had a favorable outcome, 9 (9.8%) had an unfavorable outcome, and 3 (3.3%) were not evaluated. The estimated rate of sustained treatment success was 86.4% (95% CI: 81.4%, 90.5%), 85.0% (95% CI: 78.2%, 90.4%), and 88.8% (95% CI: 80.3%, 94.5%) overall, among patients living with HIV, and among patients without HIV, respectively. Results were similar among those without a fluoroquinolone test result, with an estimated sustained treatment success rate of 85.1% (95% CI: 77.2%, 91.1%) overall, 84.1% (95% CI: 73.3%, 91.8%) among patients living with HIV, and 86.7% (73.2%, 94.9%) among patients without HIV (Table S3 in the Supporting Information). In the sensitivity analysis additionally excluding post-treatment deaths unrelated to TB, the estimated rate of sustained treatment success was 87.6% (95% CI: 82.6%, 91.5%) overall, 86.2% (95% CI: 79.5%, 91.4%) among patients living with HIV, and 89.8% (95% CI: 81.5%, 95.2%) among patients without HIV.

**Table 6:** Outcomes 18 months after treatment start among patients receiving a shortened regimen for MDR/RR-TB in Lesotho, overall and by HIV status.

| **18-month outcome** | **Overall**<br>**(N=244)**<br>**(%)** | **HIV**<br>**(N=152)**<br>**(%)** | **No HIV**<br>**(N=92)**<br>**(%)** |
| --- | --- | --- | --- |
| Favorable outcome | 204 (83.6) | 125 (82.2) | 79 (85.9) |
| Unfavorable outcome | 32 (13.1) | 22 (14.5) | 10 (10.9) |
| *Died* | 23 (9.4) | 14 (9.2) | 9 (9.8) |
| *Failed treatment* | 3 (1.2) | 3 (2.0) | 0 (0.0) |
| *LTFU during treatment* | 2 (0.8) | 2 (1.3) | 0 (0.0) |
| *Recurrence* | 4 (1.6) | 3 (2.0) | 1 (1.1) |
| Not evaluated | 8 (3.3) | 5 (3.3) | 3 (3.3) |
| *Transferred* | 4 (1.6) | 2 (1.3) | 2 (2.2) |
| *LTFU post treatment* | 4 (1.6) | 3 (2.0) | 1 (1.1) |
| Sustained treatment success<sup>1</sup>, N=236 | 204 (86.4)<br>95% CI: (81.4, 90.5) | 125 (85.0)<br>95% CI: (78.2, 90.4) | 79 (88.8)<br>95% CI: (80.3, 94.5) |

<sup>1</sup>Excludes patients who were not evaluated

Overall, the median length of post-treatment follow-up among patients with treatment success was 44 weeks (IQR 43.8-44.8). One additional death, unrelated to TB, was observed between 18 and 21 months of follow-up, for a total of 9 unfavorable outcomes during the post-treatment period (9/212 (4.2%) overall; 6/130 (4.6%) in patients living with HIV and 3/82 (3.7%) in patients without HIV). The overall rate of post-treatment recurrent TB or TB-related death was 2.2 per 1000 person-months (95% CI: 0.7, 5.2), while it was 2.2 (95% CI: 0.5, 6.4) among patients with HIV and 2.3 (95% CI: 0.3, 8.1) among patients without HIV. The incidence rate of post-treatment recurrent TB or any-cause death was 4.0 per 1000 person-months (95% CI: 1.8, 7.5) overall, 4.3 (95% CI: 1.6, 9.5) among patients with HIV, and 3.4 (95% CI: 0.7, 9.9) among patients without HIV.

## DISCUSSION

In this study, the estimated rate of sustained cure 18 months post-treatment initiation was excellent, and TB recurrence was rare. The frequency of end-of-treatment success (88.5%) surpasses that from historic cohorts of individuals treated with longer, injectable-containing regimens in Lesotho^16,17^ and is particularly striking given that the study period overlapped with the early stages of the COVID-19 pandemic. Adverse events, though common, were generally manageable and did not affect treatment success rates. This was true regardless of HIV status, suggesting that the safety and effectiveness of the nine-month regimen comprised of bedaquiline, levofloxacin, linezolid, delamanid, and clofazimine are not compromised among patients living with HIV. We expect these findings to generalize broadly to individuals with fluoroquinolone-susceptible MDR/RR-TB without prior exposure to second-line drugs; however, they may not apply to individuals who present in critical condition and/or with severe forms of extrapulmonary TB, as these patients were often excluded based on clinical discretion.

Rates of clinically relevant adverse events of special interest in this cohort, some of which are higher than in some prospective cohorts with rigorous monitoring,^18–21^ should be interpreted in the context of structured, active pharmacovigilance within routine programmatic care. Monthly clinical, laboratory, and electrocardiographic monitoring likely increased detection, relative to settings with less intensive pharmacovigilance, while enabling timely modifications, as reflected by high treatment success. The relatively higher rate of adverse events of special interest may also reflect more advanced TB disease and a generally sicker patient population. For example, anemia and body mass index less than 18.5 were highly prevalent at baseline, at 56.9% and 37.8%, respectively, and a majority of the cohort was living with HIV, many of whom had significant immune suppression. The high frequency of peripheral neuropathy of any grade was especially noteworthy. While overreporting is possible, the standardized objective and subjective screening should have minimized misclassification. In addition to advanced disease and comorbidities, other conditions common in this setting, such as nutrient deficiencies or physical exertion from occupation, may also play a role. Pinpointing causes of geographic heterogeneity in the incidence of peripheral neuropathy and other linezolid-associated adverse events remains a key area for future research.^6^

Permanent drug discontinuation due to toxicity and tolerability was common, with 61% of individuals having a drug permanently stopped, most commonly linezolid. A key benefit of the five-drug regimen is that, even if a drug is suspended, the regimen will still contain four active drugs, thereby providing greater clinical flexibility to discontinue a drug if intolerance occurs. Given the overlapping and myriad toxicities associated with currently recommended drugs for MDR/RR-TB, it is possible that five drugs may increase the frequency of adverse events, particularly in a population with comorbidities and advanced disease. However, loss to follow-up in this routine setting was rare. And, while toxicity was common, it did not appear to be appreciably worse in people living with HIV, with the possible exception of myelosuppression. The higher rate of myelosuppression among those living with HIV may reflect a direct effect of HIV infection, of opportunistic infections, and/or of concomitant medications to treat opportunistic infections.^22^ Future analyses examining which individuals are at highest risk of adverse events may inform targeted monitoring.

The high rates of treatment success despite a complex comorbidity profile and frequent linezolid modification provide reassurance of regimen resilience in a high-HIV burden, resource-constrained setting. The overall rate of sustained cure of 86.4% at 18 months is similar to those reported in trials of shortened all-oral regimens, including the BEAT-Tuberculosis trial, which reported a 86.1% treatment success rate at 76 weeks.^7^ TB-PRACTECAL reported an 89.0% success rate at 72 weeks for BPaLM,^3^ while treatment success for the three endTB trial regimens now endorsed by WHO ranged from 85.2 to 90.4%.^8^ The high rate of sustained cure at 18 months among those living with HIV (85.0%), also aligns closely with the 82.9% frequency of treatment success at 76 weeks among individuals living with HIV in the BEAT-tuberculosis trial, which included more than 200 individuals living with HIV. Success rates in the PRACTECAL and endTB trials were variable, ranging from 71.4% for BPaLM in TB-PRACTECAL^3^ to 100% for two of the endTB regimens^8^, and likely reflect the small numbers of people living with HIV in the study arms of these trials (14-17 people). Comparable effectiveness among people living with and without HIV likely reflects advances in TB/HIV care, including early antiretroviral therapy initiation, widespread dolutegravir use, and integrated service delivery. While we found no evidence that treatment effectiveness was compromised in individuals lacking a fluoroquinolone test result (i.e., due to limited DST access, reagent stockouts and additional health systems challenges associated with the early stages of the COVID-19 pandemic), this may be due to the relatively low prevalence of background fluoroquinolone resistance and the selection of patients without prior fluoroquinolone exposure; for these reasons, this finding may not generalize to other settings.

Major strengths of this study include limited missing data, the fact that all favorable outcomes were microbiologically confirmed cure, and the excellent follow-up for recurrent TB throughout the year following treatment completion. Provision of monthly food support was likely an important contributor to the high frequency of post-treatment retention. A limitation of this analysis is that although the regimen was intended to be standardized, clinical discretion was sometimes used to tailor the regimen, which resulted in variable durations of exposure to linezolid and the use of moxifloxacin, either in the initial regimen or as a replacement for levofloxacin. Rarely, a single additional drug was added for more than a month, and while the reason for this was not documented, we attribute it to the learning curve associated with clinicians adapting from longer individualized regimens to a shorter standardized regimen. A second limitation is our inability to discern between adverse events caused by a TB medication versus those caused by a non-TB medication or an underlying co-morbidity or health condition. Given that many participants received concomitant medications for conditions other than TB, the rate of adverse events may be higher than observed in other cohorts in which polypharmacy with non-TB medication occurs more rarely. Finally, these results do not inform whether individuals living with HIV, or any other group, would fare better with six months of treatment, as is done under the BEAT-Tuberculosis strategy or with the nine-months as used here.

In conclusion, a nine-month five-drug standardized regimen achieved relatively high rates of treatment success and final treatment outcomes among individuals with and without HIV infection, in spite of frequent toxicities and clinican-directed modifications to the regimen.

Future work should seek to understand the relative benefits of 6 and 9-month regimens, as well as the optimal number and composition of medications, to maximize treatment effectiveness and tolerability while minimizing toxicity.

## Data Availability

All data produced in the present study are available upon reasonable request to the authors

## Acknowledgments

SMS was supported by the National Institutes of Allergy and Infectious Diseases under Award Number T32 AI007433. MK, AL, RM, JM, DM, LM, TT, LT, EO, MC, LO, MFF, CDM, AL, KS, and MLR were supported by the National Institutes of Allergy and Infectious Diseases under Award Number R01AI46095. The funders had no role in the conceptualization, analysis, or presentation of findings of this study.

## Data sharing statement

To researchers with a methodologically sound proposal, pseudo-anonymized individual participant data that underlie the results reported in the text and tables of this article and a data dictionary may be made available upon request to the corresponding author, and execution of a data sharing agreement or alternate means that allows assurance that principles of Lesotho regulations will be met.

## Conflicts of interest

The authors report no conflicts of interest.

## Notes

### Competing Interest Statement

The authors have declared no competing interest.

