## Supplemental Information for "Nine-month all-oral regimen for MDR/RR-TB in Lesotho: safety and effectiveness by HIV"

**Supplementary Table S1.** Permanent drug discontinuation among patients who initiated a shortened all-oral regimen for multidrug-resistant or rifampin-resistant tuberculosis in Lesotho, STEM-TB cohort by LZD initiation status (N=244)

| **Received LZD**  **(N=229)** | **Drug (N who initiated the drug)** | **Discontinued**  **n (%)** |
| --- | --- | --- |
|  | Bedaquiline (N=229) | 3 (1.3) |
|  | Levofloxacin (N=221) | 14 (6.3) |
|  | Delamanid (N=229) | 7 (3.1) |
|  | Clofazimine (N=228) | 3 (1.3) |
| **Never Received LZD**  **(N=15)** | Bedaquiline (N=15) | 0 (0.0) |
|  | Levofloxacin (N=12) | 1 (8.3) |
|  | Delamanid (N=15) | 0 (0.0) |
|  | Clofazimine (N=15) | 0 (0.0) |

**Supplementary Table S2:** End-of-treatment outcomes among patients receiving a shortened regimen for MDR/RR-TB in Lesotho among 1) patients with confirmed FQ susceptibility, 2) patients missing a baseline FQ susceptibility test result

| **Confirmed FQ susceptibility** | **End of treatment outcome** | **Overall**  (N=128)  (%) | **HIV**  (N=82)  (%) | **No HIV**  (N=46)  (%) |
| --- | --- | --- | --- | --- |
|  | Cured^1^ | 115 (89.8) | 73 (89.0) | 42 (91.3) |
|  | Died | 7 (5.5) | 4 (4.9) | 3 (6.5) |
|  | Failed treatment | 3 (2.3) | 3 (3.7) | 0 (0.0) |
|  | LTFU | 1 (0.8) | 1 (1.2) | 0 (0.0) |
|  | Transferred | 2 (1.6) | 1 (1.2) | 1 (2.2) |
|  | End of treatment success^2^, N=126 | 115 (91.3)  95% CI: (84.9, 95.6) | 73 (90.1)  95% CI: (81.5, 95.6) | 42 (93.3)  95% CI: (81.7, 98.6) |
| **Missing FQ susceptibility result** | **End of treatment outcome** | **Overall**  (N=116)  (%) | **HIV**  (N=70)  (%) | **No HIV**  (N=46)  (%) |
|  | Cured^1^ | 101 (87.1) | 60 (85.7) | 41 (89.1) |
|  | Died | 12 (10.3) | 8 (11.4) | 4 (8.7) |
|  | Failed treatment | 0 (0.0) | 0 (0.0) | 0 (0.0) |
|  | LTFU | 1 (0.9) | 1 (1.4) | 0 (0.0) |
|  | Transferred | 2 (1.7) | 1 (1.4) | 1 (2.2) |
|  | End of treatment success^2^, N=114 | 101 (88.6)  95% CI: (81.3, 93.8) | 60 (87.0)  95% CI: (76.7, 93.9) | 41 (91.1)  95% CI: (78.8, 97.5) |

^1^There were no treatment completions in this cohort.

^2^Excludes patients who transferred out during treatment

**Supplementary Table S3:** Outcomes 18 months after treatment start among patients receiving a shortened regimen for MDR/RR-TB in Lesotho among 1) patients confirmed with FQ susceptibility, 2) patients missing a baseline FQ susceptibility test result

| **Confirmed FQ susceptibility** | **18-month outcome** | **Overall**  (N=128)  (%) | **HIV**  (N=82)  (%) | **No HIV**  (N=46)  (%) |
| --- | --- | --- | --- | --- |
|  | Favorable outcome | 107 (83.6) | 67 (81.7) | 40 (87.0) |
|  | Unfavorable outcome  *Died*  *Failed treatment*  *LTFU during treatment*  *Recurrence* | 15 (11.7)  8 (6.3)  3 (2.3)  1 (0.8)  3 (2.3) | 11 (13.4)  5 (6.1)  3 (3.7)  1 (1.2)  2 (2.4) | 4 (8.7  3 (6.5)  0 (0.0)  0 (0.0)  1 (2.2) |
|  | Not evaluated  *Transferred*  *LTFU post treatment* | 6 (4.7)  2 (1.6)  4 (3.1) | 4 (4.9)  1 (1.2)  3 (3.7) | 2 (4.3)  1 (2.2)  1 (2.2) |
|  | Sustained treatment success^1^, N=122 | 107 (87.7)  95% CI: (80.5, 93.0) | 67 (85.9)  95% CI: (76.2, 92.7) | 40 (90.9)  95% CI: (78.3, 97.5) |
| **Missing FQ susceptibility result** | **18-month outcome** | **Overall**  (N=116)  (%) | **HIV**  (N=70)  (%) | **No HIV**  (N=46)  (%) |
|  | Favorable outcome | 97 (83.6) | 58 (82.9) | 39 (84.8) |
|  | Unfavorable outcome  *Died*  *Failed treatment*  *LTFU during treatment*  *Recurrence* | 17 (14.7)  15 (12.9)  0 (0.0)  1 (0.9)  1 (0.9) | 11 (15.7)  9 (12.9)  0 (0.0)  1 (1.4)  1 (1.4) | 10 (10.9)  6 (13.0)  0 (0.0)  0 (0.0)  0 (0.0) |
|  | Not evaluated  *Transferred*  *LTFU post treatment* | 2 (1.7)  2 (1.7)  0 (0.0) | 1 (1.4)  1 (1.4)  0 (0.0) | 3 (3.3)  1 (2.2)  0 (0.0) |
|  | Sustained treatment success^1^, N=114 | 97 (85.1)  95% CI: (77.2, 91.1) | 58 (84.1)  95% CI: (73.3, 91.8) | 39 (86.7)  95% CI: (73.2, 94.9) |

^1^Excludes patients who transferred out during treatment or were LTFU post treatment
